# Lessons Learned from Development and Implementation of the National Surgical, Obstetric and Anaesthesia Plan in Ethiopia: a qualitative study

**DOI:** 10.1101/2023.10.06.23296670

**Authors:** Amanuel Belay, Medhanit Getachew, Abebe Bekele, Andualem Deneke Beyene, Amezene Tadesse, Justine Davies, Andrew JM Leather, Charlotte Hanlon

## Abstract

**Background:** The Saving Lives Through Safe Surgery (SaLTS) project was launched in 2016 by the Ethiopian Ministry of Health to improve access and quality of surgical and anaesthesia care throughout Ethiopia. A key action of SaLTS was to develop a National Surgical, Obstetric and Anaesthesia Plan (NSOAP). The aim of this study was to explore barriers and facilitators in the development and implementation of this plan in Ethiopia.

**Methods:** We conducted semi-structured interviews with 12 key stakeholders involved in developing and implementing the NSOAP at national and regional level in Ethiopia. Thematic analysis was performed.

**Results:** The attention given to safe surgery after the Lancet Commission on Global Surgery that reported in 2015, provided key impetus to commitment from the Ethiopian government to develop an NSOAP. Another important opportunity while drafting the plan was the engagement of relevant and motivated professionals. However, major challenges during development included a lack of consistent leadership, no prior experience to benchmark from, the extended time taken to complete the plan, the lack of exclusively dedicated staff, varying engagement, awareness and commitment of regional bureaus and poor quality of available data to inform the situation analysis. Key facilitators of implementation were ownership of the plan by the government and involvement of a multidisciplinary implementation team. However, this was undermined by poor cost estimation and inadequate commitment of financial resources, the low quality of surgical equipment and supplies, and the lack of dedicated human resources, data management systems, ongoing stakeholder engagement, and government accountability. Recommendations to increase the success of future planning and implementation included the need to properly estimate resources and financial impact, carry out a careful baseline assessment, implement checklists for monitoring and evaluation, ensure reasonable resource allocation and involve grassroots regional stakeholders from the beginning. Alongside this, countries need to have committed leadership and transparency in setting goals to bring about broad stakeholder engagement.

**Conclusion:** Ethiopia showed early commitment to strengthen surgical systems and improve surgical care. Lessons learned can help to guide and encourage other countries to develop realistic NSOAPs and support their effective implementation on the ground.

## Background

Surgical care and anaesthesia services have been recognized by the World Health Assembly (Resolution-68/15) as an important public health investment and a key component of universal health coverage [1]. Nonetheless, it has been estimated that five billion people lack access to safe, affordable surgical and anaesthesia care when needed. This disparity disproportionately affects those living in low- and middle-income countries: an estimated 93% of the population in sub-Saharan Africa does not have access to safe, affordable surgical and anaesthesia care compared to 3.6% of the population in higher-income regions [2]. In Ethiopia, a country with a population of 120 million people, there is an estimated unaddressed need for 5,000,000 surgical procedures per year [3]. To address this gap, a national plan that specifically addresses surgery is essential to ensure that surgical and anaesthesia care are fully integrated into the health system and for proper planning of care delivery, education and research [2]. Developing a national plan is an essential first step for countries to strengthen their surgical systems and improve surgical care [4].

The Saving Lives Through Safe Surgery (SaLTS) initiative was launched in 2016 by the Ethiopian Ministry of Health to improve access and quality of surgical and anaesthesia care throughout Ethiopia [3]. Through SaLTS, Ethiopia became one of the first countries to develop a National Surgical, Obstetric and Anesthesia Plan (NSOAP) and emerged as a leader in sub-Saharan Africa in prioritizing surgery as part of a national healthcare strategy [5, 6]. The Lancet Commission on Global Surgery has developed a template for the development of a national surgical plan [2], covering the following six domains aligned with the WHO health service building blocks: infrastructure, workforce, service delivery, financing, information management and governance [7]. At the time of conducting this study, six countries had developed NSOAPs, 10 countries had plans underway and 23 additional countries had shown a commitment to developing an NSOAP [6, 8].

The existence of such a national-level plan is not, however, enough in itself to ensure improved access to safe, affordable, surgical and anaesthesia care. Plans need to be implemented and we are only aware of one country (Ethiopia) that has attempted to implement an NSOAP. Although the development and early implementation of the Ethiopian NSOAP have been described previously (5), to the best of our knowledge, there has been no systematic investigation of the barriers and facilitators to the development and implementation of NSOAPs in this or any other low-resource setting. The objective of this study was to explore the NSOAP planning process and the barriers and facilitators to the development and implementation of a national surgical plan in Ethiopia.

## Methods

### Design

The study design was qualitative, nested within the surgical work package of the ASSET (HeAlth System StrEngThening in sub-Saharan Africa) National Institute of Health Research Global Health Unit [9], a health system strengthening research program working across four sub-Saharan African countries (Ethiopia, Sierra Leone, South Africa, Zimbabwe) and three platforms of care (surgery, primary care and maternal care).

### Context

The study was carried out in Ethiopia from 27^th^ May 2018 until 23^rd^ January 2020. The SaLTS initiative proposed to use a health system approach to improving surgical care through the Ethiopian Hospitals Alliance for Quality (EHAQ) platform to simultaneously tackle activities across eight ‘pillars of excellence’. These key strategic pillars are 1) Leadership, management, and governance: a nationally scaled surgical leadership and mentorship program, 2) Infrastructure: operating room construction and oxygen delivery plan, 3) Supplies and logistics: a national essential surgical procedure and equipment list, 4) Human resource development: a Surgical Workforce Expansion Plan and Anaesthesia National Roadmap, 5) Advocacy and partnership: strong Ministry of Health partnerships with international organizations, including General Electric Foundation’s safe Surgery 2020 initiative, 6) Innovation: facility-driven identification of problems and solutions, 7) Quality of surgical and anaesthesia care service delivery: a national peri-operative guideline and WHO Surgical Safety Checklist implementation, and 8) Monitoring and evaluation: a comprehensive plan for short-term and long-term assessment of surgical quality and capacity [3].

### Participants

Informants were purposively selected based on being key stakeholders involved at the national or regional level of plan development and implementation. They included policy-makers, planners, surgeons in leadership positions, individuals with a variety of responsibilities at NGOs and government level consultants.

### Data collection

A topic guide was developed for in-depth interviews based upon the authors’ knowledge of the field and the literature. The interview guide was developed to explore the following topics: the process of developing the plan, main steps taken during the drafting of the plan, opportunities and barriers during the development process, key players in the development process, opportunities and barriers during the implementation of the plan and lessons learned/recommendations for future planning.

The topic guide was first developed in English and translated into Amharic. One experienced interviewer, fluent in Amharic did all the interviews in Amharic. Interviews were audio-recorded. Eleven interviews took place in the respondents’ office and one was done virtually via the zoom platform.

### Analysis

All interviews were transcribed in Amharic, translated into English, and uploaded to OpenCode 4.02 software to facilitate analysis. Thematic analysis was performed. Two members of the research team (AB, MG) read and re-read transcripts to familiarize themselves with the data and independently carried out line-by-line coding of two randomly selected transcripts. AB and MG met twice to compare and refine codes and coding. They developed a common codebook through a discussion of each code definition and assignment. CH reviewed the codebook with AB and MG to identify areas of overlap, inconsistency or possible gaps. AB and MG then independently coded the remaining transcripts based on the codebook. Similar codes were grouped to form themes, and these were compared back to the data and iteratively improved until consensus was reached. Quotes from a range of participants were selected to illustrate these themes.

### Ethical approval

Ethical clearance was obtained from the Institutional Review Board (IRB) of the College of Health Sciences of Addis Ababa University (protocol number 026/18/PSY) and King’s College London (protocol number HR-17/18-6144). Written informed consent was obtained from each participant. Confidentiality was ensured by anonymising identifying information and assigning a unique ID code to all interviews.

## Results

Twelve respondents participated, including respondents from the Ministry of Health, a Regional Health Bureau and implementing NGOs. See Table 1 for characteristics. Ten participants were trained as medical doctors, one was a nurse and one had a public health background.

**Table 1.** Socio-demographic characteristics of participants.

| Code | Position | Involvement in the Saving Lives Through Safe Surgery (SaLTS) initiative |
| --- | --- | --- |
| ID 01 | Ministry of Health | Planning/design and implementation |
| ID 02 | Ministry of Health | Planning/design only |
| ID 03 | Implementing NGO | Planning/design and implementation |
| ID 04 | Implementing NGO | Planning/design |
| ID 05 | Implementing NGO | Late stage of the planning/design |
| ID 06 | Surgical clinical expert | Planning/design |
| ID 07 | Regional Health Bureau | Piloting stage |
| ID 08 | Surgical clinical expert | Planning/design and implementation |
| ID 09 | Surgical clinical expert | Piloting stage |
| ID 10 | Surgical clinical expert | Piloting |
| ID 11 | Ministry of Health | Planning/design and implementation |
| ID 12 | Surgical clinical expert | Planning/design and implementation |

### Key steps in drafting and developing the NSOAP

Safe surgery as a problem area needing a response started to be given attention in Ethiopia following the publication of the Lancet Commission on Global Surgery in 2015 (Fig 1). The adoption of emergency and essential surgical care and anaesthesia as a component of universal health coverage at the May 2015 World Health Assembly (WHA) was also perceived by respondents as providing important impetus for the Ethiopian Ministry of Health (MoH) to prioritize surgical care and initiate development of the NSOAP.

**Figure 1.**
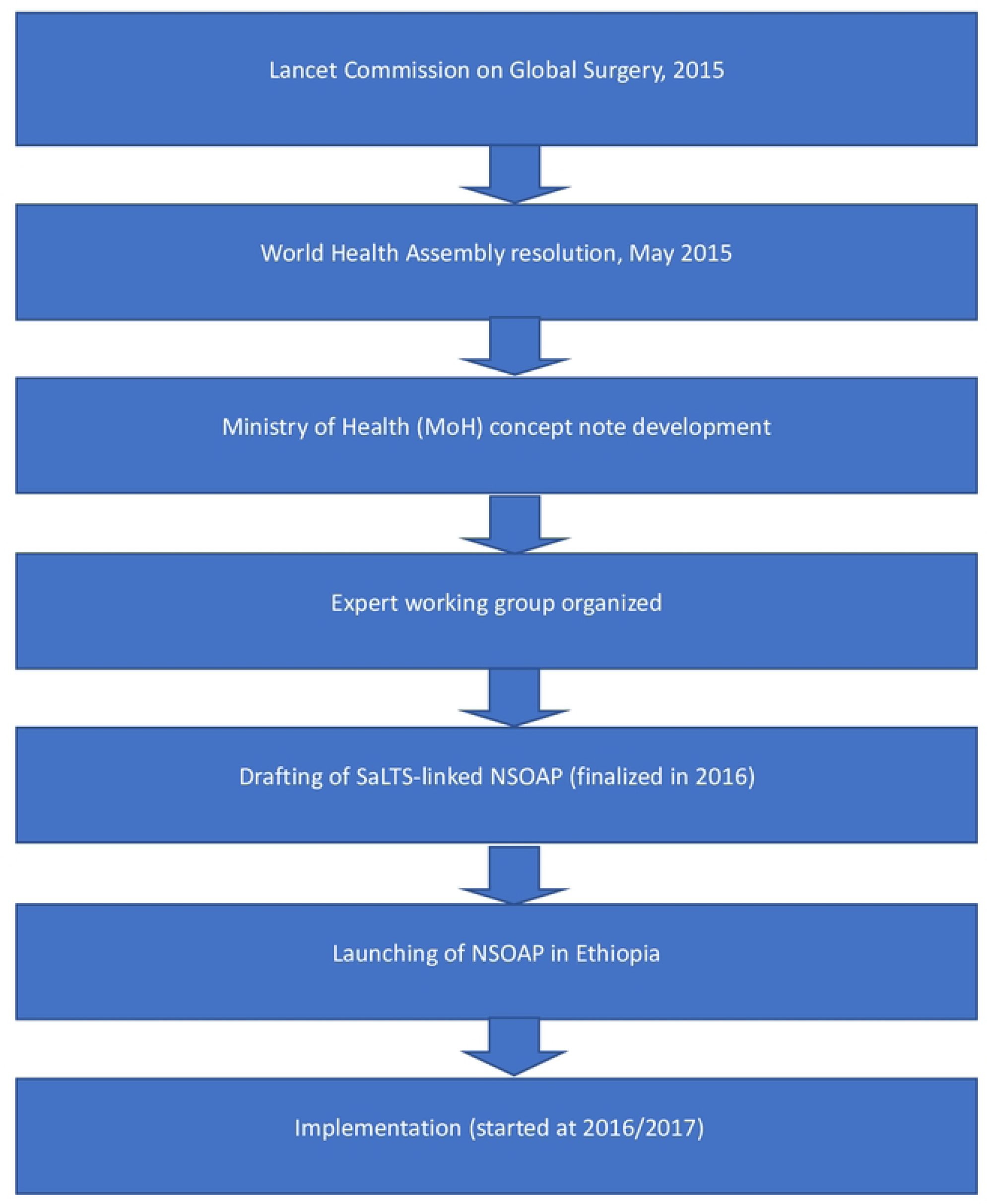
Key steps in the development of the Ethiopian NSOAP.

The next key step was the MoH engaging with various professional institutions and stakeholders. The idea for SaLTS was presented to senior surgeons, anaesthetists and obstetricians in Ethiopia. Under the medical services directorate of MoH, a team of professionals was formed to provide technical support and develop a concept note for SaLTS. The team members were identified through the Ethiopian Medical Association and Addis Ababa University. To prioritize surgical interventions that are cost-effective for scaling up, an “Ethiopian essential service package for surgery” was developed. During the development process of this package, in addition to using the documents prepared by World Bank and WHO as a baseline, input was obtained from a multi-professional team drawn from multiple different surgical disciplines (including Ear, Nose and Throat, and dental).

> *“… engagement was a critical part but during the engagement process, it was necessary to engage the responsible professionals. Therefore, senior professionals have engaged not only once but also repeatedly … and the other thing we shouldn’t have to underestimate is, technical support, which is a very important thing. There were a team of professionals that came from different areas who were dedicated to support and advise the ministry of health.” (ID 03)*

Another important step in developing the surgical plan was reported to be the time taken to conduct a situational analysis. After a concept note was developed for funding, the situation analysis was carried out. The situation analysis focused on the current status of infrastructure, human resources, equipment in different settings and surgical and anaesthesia services that were being provided across Ethiopia. The data from this situation analysis was used to identify gaps and helped the team to develop the eight pillars for the Plan.

> *“…… to some extent situational analysis was attempted. This situational analysis, when it is done, the pillars, for instance, on HR [human resources], how many surgeons are there, how many obstetricians do we have, how many ophthalmologists do we have, where are they and the like, these kinds of exercise was done on the analysis. Then, starting from the situational analysis, after we identify gaps, those eight pillars were made to stand out.” (ID 01)*

The planning and implementation of some pillars of the NSOAP were described as being undertaken in parallel. Along with the planning process, anaesthesia, Intensive Care Unit and Operating Room equipment that was needed for implementation was purchased. In addition, residency programmes for training of anaesthetists, surgeons, obstetricians and ophthalmologists were expanded to strengthen the available workforce. Respondents emphasized the importance of piloting aspects of the plan. As part of the implementation, a program on human capacity building, which aimed to strengthen leadership and innovation for surgical and anaesthesia care was piloted in five hospitals located in two regions. The experience from this pilot study was then incorporated into the national plan (SaLTS).

> *“… For learning purposes, as the two regions were ready at the time, we conducted pilot studies in five hospitals in Amhara and Tigray regions. Then we took the experience we found and we have tried to incorporate it as input in the SaLTs document.” (ID 08)*

### Challenges and opportunities in developing the NSOAP

Many of the respondents spoke of the opportunity that arose due to the global attention given to safe surgery and the resulting recognition of surgery as an essential component of care in Ethiopia.

> *“The Lancet Commission publication is a landmark publication on the role of surgery as an essential [part of] universal care. After that, they [Ministry of Health officials] believed that there is a need to have one strategic document for Ethiopia too. So, the minister who was there at that time believed in that and started the initiative and developed the plan together with the experts…” (ID 05)*

The resulting commitments from champions for surgical care in the MoH and the funding made available by various partners were mentioned as important opportunities. The strong engagement of stakeholders, especially professionals and professional societies, (e.g. the Surgical Society of Ethiopia) was also seen as vital to drive the process forwards.

An important challenge was that this was the first time an NSOAP had been developed in a low-income country. There was no prior experience or benchmark, so this made the process difficult.

> *“…at that time, when we did this, no other country had developed their surgical plan so, this was I can see the first so, when you are the first, again the problem is there is no one you can benchmark or refer to, there was no one we could do that with, so it was not easy …” (ID 01)*

Respondents noted that a lack of consistent leadership and management of the process of developing and implementing the plan was one of the main challenges. As the development process went on, the policymakers were perceived not to have the required understanding and commitment to lead. It was also noted that the leadership did not allow active participation of all key stakeholders or engagement was late, when decisions had already been made. Inconsistent leadership was described by respondents, exacerbated by a high turnover of leadership within the MoH. Friction about who should lead was also present.

> *“First, the leadership at the ministry of health was good in principle. But there was a turnover of the leadership team. There was inability to make priorities most of the times. There was sometimes no strong tendency from the Ministry of Health side to bring all together and communicate in a clear manner and take things forward. So, from the ministry of health side, a deficiency was observed.” (ID 08)*

Not including a budget allocation plan due to the uncertainties of how much it would cost was mentioned as the main limitation of the plan.

> *“But I think the main limitation of the plan was, budget allocation was not done…the budget allocation was not seen on the plan. But the plan has a big impact on the budget…I think it is because if the budget is predetermined it might be very high or it might be very low …I think that is why the budget is not calculated. We have tried to draft the budget on another document but if you see the document now, you may not find the exact amount of money.” (ID 06)*

The lengthy time for the planning process was emphasized by respondents as the other main challenge. In total, the planning process took about nine months. No staff members were exclusively dedicated to the task; rather the team had to drop other activities, like delivery of surgical care, to engage in the planning process. There were competing priorities of the surgeons that did not allow them to provide timely review and feedback to inform the planning process.

> *“The other challenge I would think is you would have competitive priority and like the team does not do this full time, many of them are physicians, they have to go to their clinic, they have to go to their patients …“ (SALT 10)*

> *“It is as I told you, as the individual didn’t engage, it might not be timely reviewed and things might not go by the pace you thought, and there were competing priorities. Due to these reasons, it was late for a certain time.” (SALT 04)*

Furthermore, limited data availability and poor quality of the available baseline data was a barrier to progress. Basic information on essential service packages and the number of practitioners in the country were not readily accessible.

> *“It is difficult to get quality data in our country. We don’t even know the burden of surgical disease; it is not easy to know the number of practitioners in each field. If you want to know how many anaesthetists are actively practising in Ethiopia, you wouldn’t find the number. So, it is difficult to find such type of background data…it is difficult to find quality data. Thus, if you are thinking to develop the plan based on that data, it will be difficult.” (ID 05)*

Lack of reporting from the devolved regional health administrations, reflecting both a lack of awareness and involvement, was one of the reasons identified for limited data sources. The variable understanding, commitment and willingness of regional health bureaus to invest in and lead the implementation of the plan was perceived to be an important barrier.

### Implementation Challenges and facilitators (current and foreseen)

Although respondents had some criticisms of the MoH leadership team during the development of the plan, the leadership at all levels and ownership of the program by the government (MoH) was seen as vital for actually getting the plan implemented.

> *“… At its inception, it was Dr Kesete’s (the minister at the time) idea, not anyone else’s. If we give credit, it should be for him. He identified the funding and he had the idea that the focus area should be on surgery and the country should focus on that. He was consulting others about this idea… We have repeatedly discussed how we should design the project. Then his project was written by engaging others in the ministry and surgeons outside the ministry, then when a partnership was started with GE Foundation, Jhpiego was involved and then Harvard was engaged, it meant that these all were actively engaged.” (ID 12)*

Another important facilitator was bringing together professionals from relevant professional associations. The involvement of a multidisciplinary implementation team of surgeons from the Ethiopian Surgical Society, partner NGOs and other professionals (especially anaesthetists) was seen as a strength.

Respondents also mentioned monitoring and evaluation, presentation of findings to higher officials and stakeholders, follow up, accountability, decentralization of the work to regional health offices, better resources and infrastructure and detailed planning as important implementation facilitators.

Nonetheless, the success of implementation was greatly affected by limitations in financial resources and surgical supplies. Problems with the systems for purchasing and maintenance of equipment and supplies, as well as low quality of the procured materials, held back implementation.

> *“Supply issues, the sustainability of consumable supplies, anaesthesia and other surgical equipment’s whether it is consumable or non-consumable, when we think of the sustained availability of these supplies, not only in our country but also worldwide, some of the supplies are not accessible, they are not affordable as well.” (ID 11)*

Respondents said that the cost estimation exercise was not sufficiently thorough or accurate which had consequences for the execution of the plan. In addition, different implementation activities were funded by different donors but the fund was only available for a few selected hospitals, making the broader execution of the plan nationwide unfeasible.

As with the development process, changes in the MoH leadership, the lack of a dedicated lead person, limited resources and high turnover rates of professionals at the implementation sites undermined implementation efforts.

> *“What challenged the programme was the leadership change at the Ministry of Health. A project which used to be seen greatly, a project which reports used to be requested weekly by the ministry, which was a regular meeting agenda, a project raised by the minister whenever he travels abroad, it all went down to a cliff as soon as a new minister is appointed,” (ID 12)*

In addition, the prominent involvement of NGOs at the implementation stage made the health professionals consider SaLTS as additional work as opposed to part of the routine work.

> *“… the idea of “donor’s job” is usually “NGO’s job” and that will be a barrier. Thus, as much as possible we need to take that out of our minds. We need to convince them as this is the ministry’s job and as it is part of our routine work. Otherwise, it is going to be a barrier. When you are going to do some tasks, if it is NGO’s job, people have a different attitude towards that and they might ask for additional payment. So, such kinds of things might happen more than you realize. Because, they may say like, “this is NGO’s job and it is not clear to understand” … such type of feelings might be a barrier.” (ID 05)*

Respondents mentioned that the political instability in the country that began just before implementation started was also a huge challenge.

> *“… right at our entry point, the country went into chaos. Those people that we trained on SaLTS, they back out, that means they have resigned in regions that means, things that were there, all people and those who took the orientation and training, those who took the skill, the people who took it, overall, given the situation that was there in the country, the economy inflation and the situation in the country, they resigned, so, when you go and ask them what SaLTS is they talk to you about the salt we put in foods, not this Safe lives through safe surgery.” (ID 02)*

The lack of a data management system, ongoing stakeholder engagement and government accountability were also raised as challenges to implementation. A few participants stated that the programme did not consider how surgery fits with other priority programs or the affordability for the community.

### Lessons learned in developing and implementing an NSOAP

There were several recommendations by respondents for other countries that intend to develop and implement national surgical plans. Framing safe surgery as a public health problem and ensuring committed leadership at all levels were the key points mentioned by respondents. It was said that professionals in the sector should stay motivated and give national planning and implementation high priority. To maintain motivation, the MoH or other stakeholders that own the process in the country should decentralize the work to the grassroots level and encourage leadership at all levels.

> *“The first thing is to make either the political leaders or technical leaders own the program. It is important to make them take that as their own job. If we can do this, the leadership at each level will have a responsibility and accountability…and they will allocate resources for the program and they will also evaluate whether the resource is being used properly or not. Therefore, it is important to do this starting from the federal level to the woreda level.” (ID 06)*

Transparency in setting the national goals and commitment by the governing body would allow the stakeholders at different levels to have similar understandings and strengthen their commitment.

> *“The major thing is to have a clear plan. There has to be a clear plan, the plan should clearly state the implementation process and responsibilities. And there needs to be monitoring and evaluation to follow evaluate the performance of the implementation. But there need to be standards to be followed while doing this.” (ID 11)*

Another important recommendation was the need for a community baseline assessment to develop a contextualized plan. Such an assessment should include situation analyses of resources and financial impacts of surgical conditions to make the case for the return on investment for making surgical care available.

> *“It is good to conduct a situational analysis in the whole country. It is important to know how many hospitals are available, the level of those hospitals, how many health centres there are, after that it is also necessary to assess the availability of the human resource. They need to think about the availability of human resources that can implement the program.” (ID 06)*

Furthermore, it was recommended that the planning process should be done step by step, ensuring that all required tasks are completed promptly. This will allow partners to take part in steps that require their input. This will additionally allow the process to be comprehensive, for instance, considering all types of surgery, and other tasks needed when planning. Finally having checklists and standards for monitoring and evaluation of the plan and setting ethical standards were highlighted as key points to consider when drafting such a plan.

Changes that the respondents suggested in the planning and design process included engagement of stakeholders at all levels (mainly regional bureau staff) from the beginning to the end. This engagement should be done in the form of teamwork, where both leaders and team members are equally informed and committed. The rationale for this approach was that it would maintain commitment despite a change in the leadership. One respondent suggested that the development of an activity-based strategic plan before a national surgical plan would have given better direction to implementation activities.

> *“The first thing is the leadership commitment has main role. We have to make commitment at Ministry of Health and regional level by all aspects and engage professional societies since they are the one which implement it. So, their engagement have main role.“ (ID 04)*

It was suggested that the pillars of SaLTS should be refined using local experience; for example, to add one pillar on social mobilization and to prioritize fewer pillars to give more focus on details. Developing the plan at the country’s regional level and consolidating centrally at a later stage was also recommended as an alternative approach to top-down planning and implementation. Regional planning would need to be supported by adequate budget, facilitation and human resources.

> *“I believe that it is also necessary to work on social mobilization and it might be necessary to add one pillar which can address the social mobilization part. Because it is important to take SALT program from the health professionals and ministry of health and give the ownership to the community.” (ID 06)*

## Discussion

This paper explored the processes, facilitators and barriers of development and implementation of an NSOAP in Ethiopia.

The NSOAP was the first of its kind to be developed and implemented in Ethiopia. A major opportunity that supported the development and implementation process was the recognition given to safe surgery and anaesthesia globally, which gave impetus to national recognition of this neglected area of healthcare. The involvement of Ethiopian global surgery experts in the global surgical movement is likely to have facilitated this translation from global to national relevance. There are increasing calls for global health initiatives to be led from the global south [10]. Strong LMIC leadership is likely to increase buy-in by national governments while ensuring contextual relevance and alignment with priorities.

Key identified challenges resulted from the lack of consistent leadership at the highest level, full commitment from stakeholders and the availability of limited resources. The high-level political commitment by the Ministry of Health was the first key step in the development of the NSOAP. Strong ownership by the government and the involvement of expert working groups that included key individuals and organizations involved in surgery and anaesthesia was the next important step in the development of the plan. The Ethiopian Ministry of Health has a strong track record of exercising country ownership of health development [11]. Thus, MoH tends to coordinate resources from partners and donors so that they are directed towards the priorities of the country and avoid fragmentation from vertical planning or undue influence of external agendas. Closely related to the question of political commitment is the imperative of ensuring strong and continuous leadership which, our findings indicate was one of the main challenges in both the development and the implementation of this national plan. Inconsistent or inadequate leadership has been identified as a barrier to implementation of national plans for non-surgical initiatives in other LMICs as well [12]. Even though there was a strong initial engagement of stakeholders in developing the NSOAP, there was a lack of sustained engagement in the implementation of the plan. Systematically identifying and involving relevant stakeholders in developing such a plan and continued engagement of all stakeholders at implementation is necessary [13].

A major implementation challenge for the Ethiopia national surgical plan was the inadequacy of available funding. Limited financial resources are a key challenge in implementing health plans in other countries [14, 15]. Not having a budget allocation plan for such a country-wide programme undermines implementation efforts, even when significant financial support is available from partners. Lessons learned from the description of developing and planning to implement the national surgical plan in Tanzania showed that the funding available was sufficient but required dedicated staff to properly manage the programme and allocate funds for the planned activities [16]. This lack of dedicated staff was similarly pointed out as a challenge in our study. Findings from early implementation of the Ethiopia NSOAP had shown that attracting additional financial resources was a challenge [5]. The importance of ensuring sustainable funding for such programs is also backed by the literature [17]. A vital strategy to guaranteeing a budget line for NSOAP implementation is integration of an NSOAP into a country’s national health strategy and involvement of the Ministry of Finance from the earliest stages of NSOAP development [4, 18].

Identifying and utilization of domestic funding to be supplemented with funding from international partners is also important [4, 18–20]. Studies show that essential surgical procedures are among the most cost-effective of all health interventions and that investment in surgical services in LMICs is affordable [2, 21]. Without scale-up of surgical care, surgical conditions are estimated to result in losses of up to 2.5% potential annual GDP in LMICs by 2030 [22], making the investment in surgical care compelling from an economic point of view alone.

The designing of the Ethiopian NSOAP could have been more effective if it was based on more detailed and better-quality data, as well as more diverse data sources. A baseline situational assessment could also add value to the quality of the plan and inform implementation indicators and targets [4, 7]. Data points for constructing these indicators and the situation should be established from regular collection and reporting of standard metrics [23]. A strong monitoring and evaluation plan is essential for successful NSOAP implementation [24]. Poor and limited data along with inadequate data management infrastructure and capacity were highlighted by our respondents. This was also mentioned in the literature as a key barrier to effective monitoring and evaluation [25, 26].

A limitation of this study is that the only source of our findings were individuals who participated in the planning, designing and piloting of the surgical plan from the ministry of health and NGOs. Individuals working at the regional level and health facilities did not participate in the study and so we may have missed important insights on the implementation of SaLTs. Furthermore, the voices of patients and advocacy groups were not included. It is important for future researchers to ensure that the experiences and priorities of more diverse stakeholders are explored.

In conclusion, country-led planning and the involvement of motivated professionals has been critical for strengthening the surgical and anaesthesia systems and improving surgical care in Ethiopia. Lessons learned from the experience of developing and implementing a surgical plan in Ethiopia can improve ongoing implementation, while also providing important insights to guide other countries to achieve realistic and effective surgical system change.

## Data Availability

Data cannot be shared publicly because the interview transcripts could lead to identification of individual participants. Data are available from the authors for researchers who meet the criteria for access to confidential data.

## Author contributions

AL, JD and AB conceptualised the overarching research goals and aims, and contributed to funding acquisition. AL, JD, AB, AD and CH contributed to developing the methodology. MG conducted the interviews. AB and MG led data analysis, supervised by CH. AB and MG prepared the first draft. All authors contributed to interpretation of findings, reviewing and editing drafts to produce the final manuscript. All authors approved the final submitted version of the manuscript.

## Acknowledgments

The research underpinning the findings presented in this paper was funded by the National Institute for Health and Care Research (NIHR) Global Health Research Unit on Health System Strengthening in Sub-Saharan Africa (ASSET), King’s College London (GHRU 16/136/54) using UK aid from the UK Government. Charlotte Hanlon (CH) receives support through an NIHR RIGHT grant (NIHR200842) and an NIHR global health research group on homelessness and mental health in Africa (HOPE; NIHR134325). The views expressed in this publication are those of the authors and not necessarily those of the NHS, the National Institute for Health and Care Research or the Department of Health and Social Care, England. CH is also funded by the Wellcome Trust through grants 222154/Z20/Z (SCOPE) and 223615/Z/21/Z (PROMISE).

For the purposes of open access, the author has applied a Creative Commons Attribution (CC BY) licence to any Accepted Author Manuscript version arising from this submission.

